# DIAGNOSTIC ACCURACY OF NON-INVASIVE DETECTION OF SARS-COV-2 INFECTION BY CANINE OLFACTION

**DOI:** 10.1101/2022.03.07.22271219

**Authors:** Dominique Grandjean, Caroline Elie, Capucine Gallet, Clotilde Julien, Vinciane Roger, Loïc Desquilbet, Guillaume Alvergnat, Séverine Delarue, Audrey Gabassi, Marine Minier, Laure Choupeaux, Solen Kerneis, Constance Delaugerre, Jérôme Le Goff, Jean-Marc Treluyer

## Abstract

**BACKGROUND:** Throughout the COVID-19 pandemic, testing individuals remains a key action. One approach to rapid testing is to consider the olfactory capacities of trained detection dogs.

**METHODS:** Prospective cohort study in two community COVID-19 screening centers. Two nasopharyngeal swabs (NPS), one saliva and one sweat samples were simultaneously collected. The dog handlers (and the dogs…) were blinded with regards to the Covid status. The diagnostic accuracy of non-invasive detection of SARS-CoV-2 infection by canine olfaction was assessed as compared to nasopharyngeal RT-PCR as the reference standard, saliva RT-PCR and nasopharyngeal antigen testing.

**RESULTS:** 335 ambulatory adults (143 symptomatic and 192 asymptomatic) were included. Overall, 109/335 participants tested positive on nasopharyngeal RT-PCR either in symptomatic (78/143) or in asymptomatic participants (31/192). The overall sensitivity of canine detection was 97% (95% CI, 92 to 99) and even reached 100% (95% CI, 89 to 100) in asymptomatic individuals compared to NPS RT-PCR. The specificity was 91% (95% CI, 72 to 91), reaching 94% (95% CI, 90 to 97) for asymptomatic individuals. The sensitivity of canine detection was higher than that of nasopharyngeal antigen testing (97% CI: 91 to 99 versus 84% CI: 74 to 90, p=0.006), but the specificity was lower (90% CI: 84 to 95 versus 97% CI: 93 to 99, p=0.016).

**CONCLUSIONS:** Non-invasive detection of SARS-CoV-2 infection by canine olfaction could be one alternative to NPS RT-PCR when it is necessary to obtain a result very quickly according to the same indications as antigenic tests in the context of mass screening.

## INTRODUCTION

Facing the COVID-19 crisis, early and efficient screenings are needed to limit the spread of the virus, allowing for the prompt isolation of positive individuals, a key action in this constant fight. Presently, most ongoing diagnostic COVID-19 testing involves nasopharyngeal sampling for RT-PCR, nasopharyngeal point-of-care antigen testing or saliva RT-PCR to identify the pathogen. Nasopharyngeal sampling for RT-PCR is the reference test but has the drawback of invasiveness and discomfort (nasopharyngeal sampling) and/or delay (RT-PCR) in obtaining the result.

Volatile organic compounds (VOC) have the potential to become a revolutionary and non-invasive approach to medical diagnostic in humans for conditions like cancer and degenerative or infectious diseases. Aksenov [1,2] studied the VOCs produced by cultures of B-lymphocytes infected by three influenza viruses: avian H9N2, avian H6N2 and human H1N1. The families of collected VOCs turned out to be unique and specific to each viral subtype. The authors concluded that the minor alterations induced by the virus on the cell’s genome expression led to a specific change in the production of VOCs in the cellular metabolism. Recently, Abd El Qader [3] showed the specificity of bacterial or viral species in the VOCs produced by infected cell cultures. Schivo [4] also identified a specific volatilome on airway cells infected by a rhinovirus. Canine olfactory detection capacities have been utilized for years in police enforcement to detect narcotics or forensic remains, explosives, bank notes, for human search and rescue missions and even to locate landmines.

Our hypothesis, the object of our previous proof of concept [5], was based on the potential excretion of specific VOCs in the sweat, induced by SARS-CoV-2 cellular actions or replications generating VOCs that the dogs can detect.

The objective of this prospective blinded multicenter study was to compare the diagnostic accuracy of the detection of SARS-CoV-2 infection by canine olfaction with the current reference standard (nasopharyngeal RT-PCR) and two alternate diagnostic strategies (nasopharyngeal antigenic test and saliva RT-PCR) in community testing centers.

## MATERIALS AND METHODS

This research is part of the “SALICOV-APHP” study (Evaluation of a strategy of SARS-CoV-2 infection testing on a general population, based on the utilization of new detection or diagnostic orientation approaches), promoted by Assistance Publique-Hôpitaux de Paris, France. The SALICOV-APHP study was approved the Protection of Persons Committee (CPP) Ile-de France III (number 3840-NI) and is registered on clinicaltrial.gov (NCT04578509). The protocol was approved for the dogs by the committee on the ethics of animal experiments of the Ecole Nationale Vétérinaire d’Alfort. All research procedures were employed in accordance with the relevant guidelines and regulations.

### Recruitment of participants

Participants were recruited from two COVID screening centers in Paris piloted by APHP. Informed consent was obtained from all subjects. Eligible persons received detailed oral and written information[6]. Participants were prospectively enrolled if they were not opposed to participating in the study. The following data were collected: age, gender, medical background, current symptoms (temperature higher than 37.8°C, chills, cough, rhinorrhea, muscular pain, loss of olfaction or taste, persisting headache, severe asthenia, etc.) and their onset date, alcohol/coffee/food/tobacco consumption or tooth cleaning within 2 hours and 24 hours prior to the test.

### Samples

Sweat samples were collected by dedicated APHP sampling teams by asking the participants to place two sterile surgical compresses under their armpits for 2 min. Samples were stored in sterile medical anti-UV glass containers, disinfected by the sampler’s helper, anonymously coded, then placed into a second plastic envelope. Individual anonymous data were registered by APHP staff for each coded sample. All samples were transferred within the day of collection from the sampling site to the testing site in coolers that were cleaned and disinfected with a 10% aqueous acetone solution after each use. All samples were stored until they were to be sniffed by the dogs in a minus 20°C freezer and were never manipulated without disposable surgical gloves to prevent contamination. Nasopharyngeal samples (NPS) were collected by a trained nurse, and participants were asked to self-collect the saliva sample after swishing saliva in their mouths for 30 seconds [6].

### Canine resources

The dogs involved in this study belong to French fire departments (Service départemental d’Incendie et de Secours – SDIS of Yvelines and Oise) and to the Ministry of the Interior of the United Arab Emirates (provided on the occasion to increase the number of operational dogs involved).

They all were trained in the same way according to the training protocol we developed in a previous publication, with line-ups of positive and negative olfaction cones based on positive reinforcement (toys) and validated similarly for their individual sensitivities and specificities [5].

The welfare of the dogs was fully respected, with toy rewards and a total absence of work-induced physical or mental fatigue. This study was carried out in strict accordance with the recommendations published in the guide for the care and use of animals edited by French law (articles R214-87 to R214-137 of the French Rural Code).

### Testing protocol

The testing sessions took place in a dedicated room (Figures 1) in the Alfort School of Veterinary Medicine. A line-up consisting of 10 olfaction cones was placed in the room. During trials, all the cones contained the unfamiliar samples, so neither the dog handlers, the data recorder nor any of the individuals present knew anything about the positivity/negativity of the samples. Once the dogs had performed the line-up, the cones and the background were cleaned with high pressure vapor, new samples were placed and a new trial cycle could start. For each new sample placement, the person in charge had to wear new disposable gloves (of the same brand during the entire period of testing sessions) and a mask in order not to contaminate the olfactory environment. Each line-up was sniffed by at least two dogs in order to mimic a real operational situation.

**Figure 1A:**
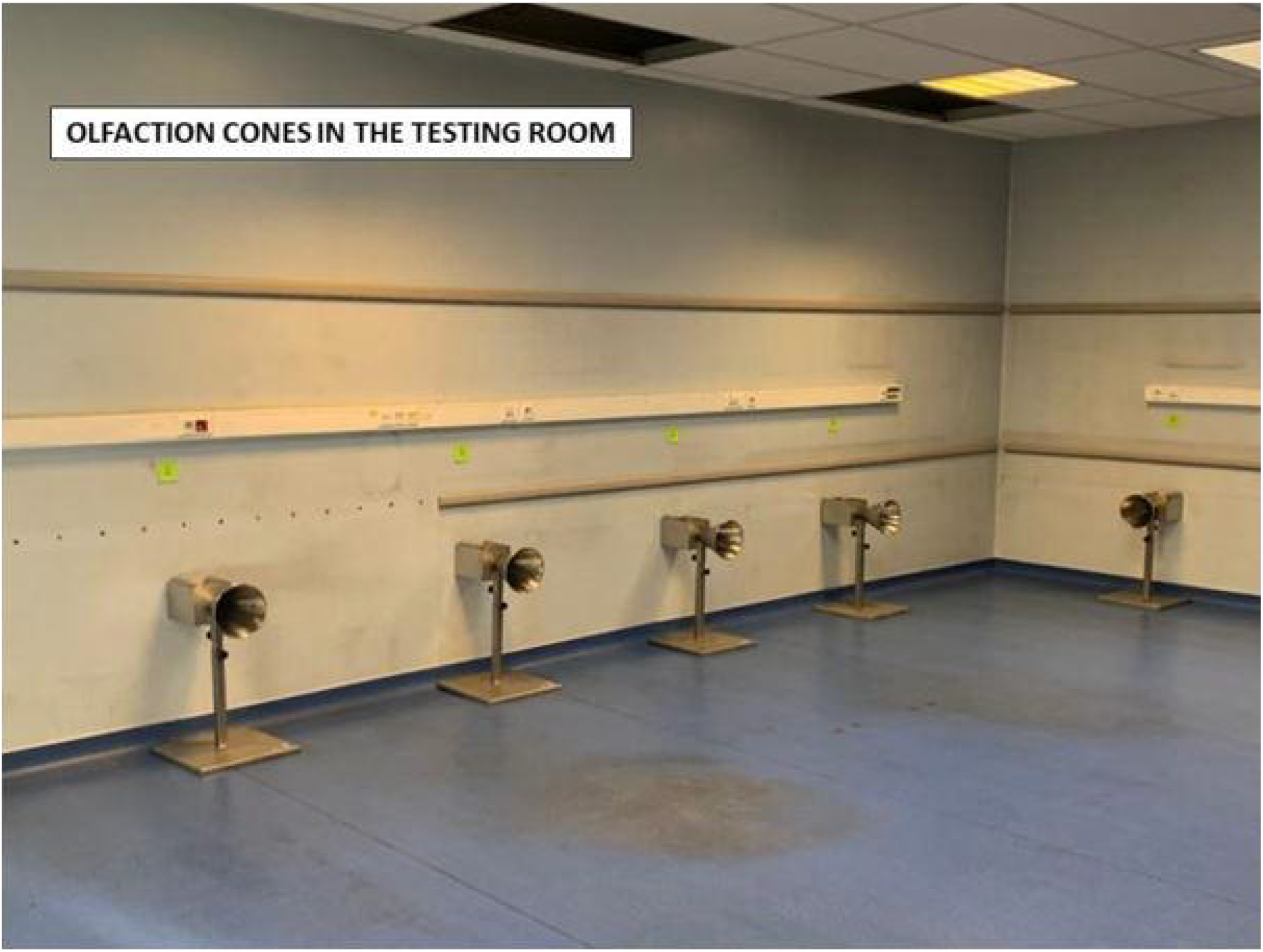
Testing room with its olfaction cones

**Figure 1B:**
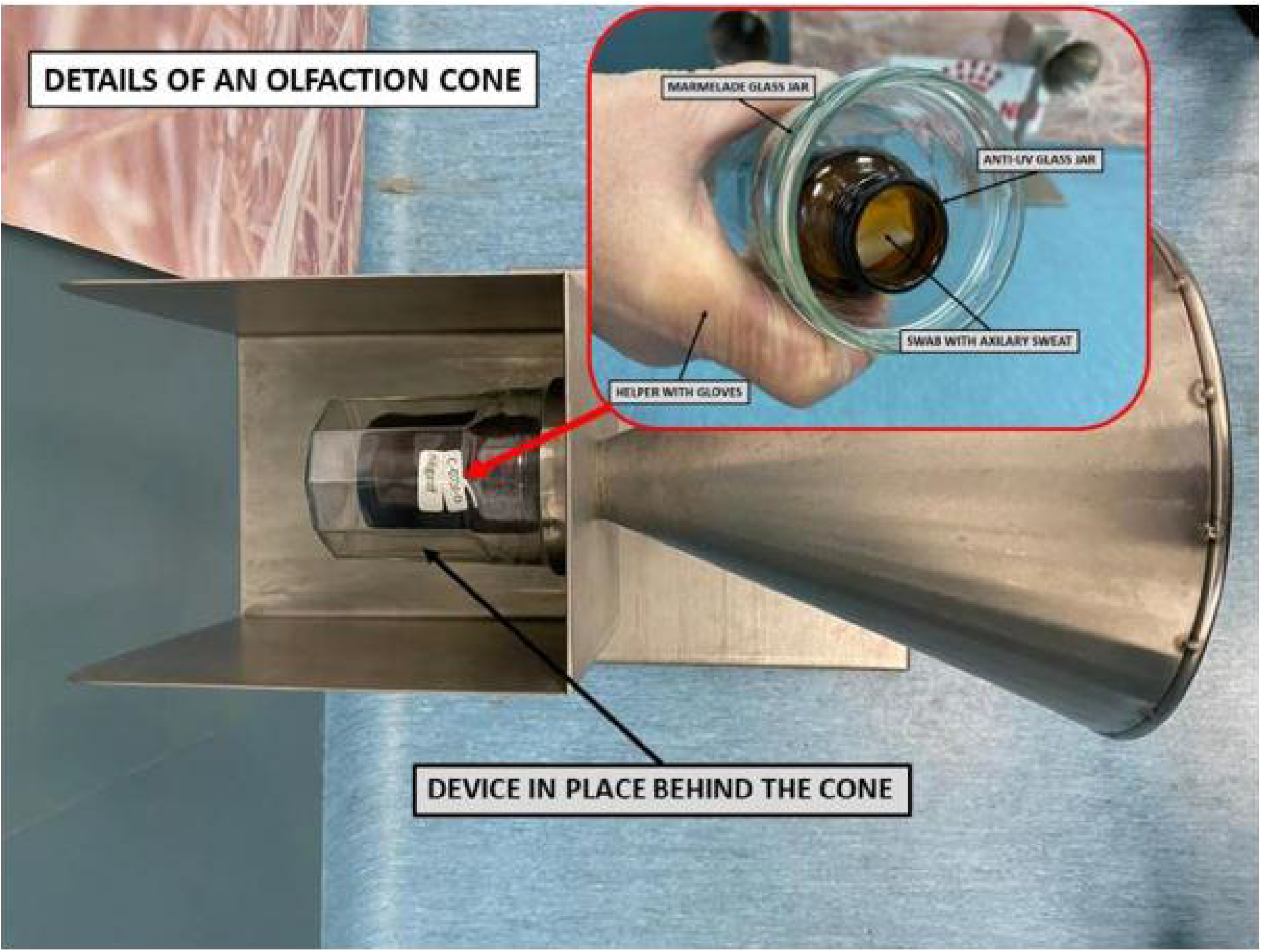
Details of an olfaction cone, with a double-protected sample and no possibility of direct contact with the dog

**Figure 1C:**
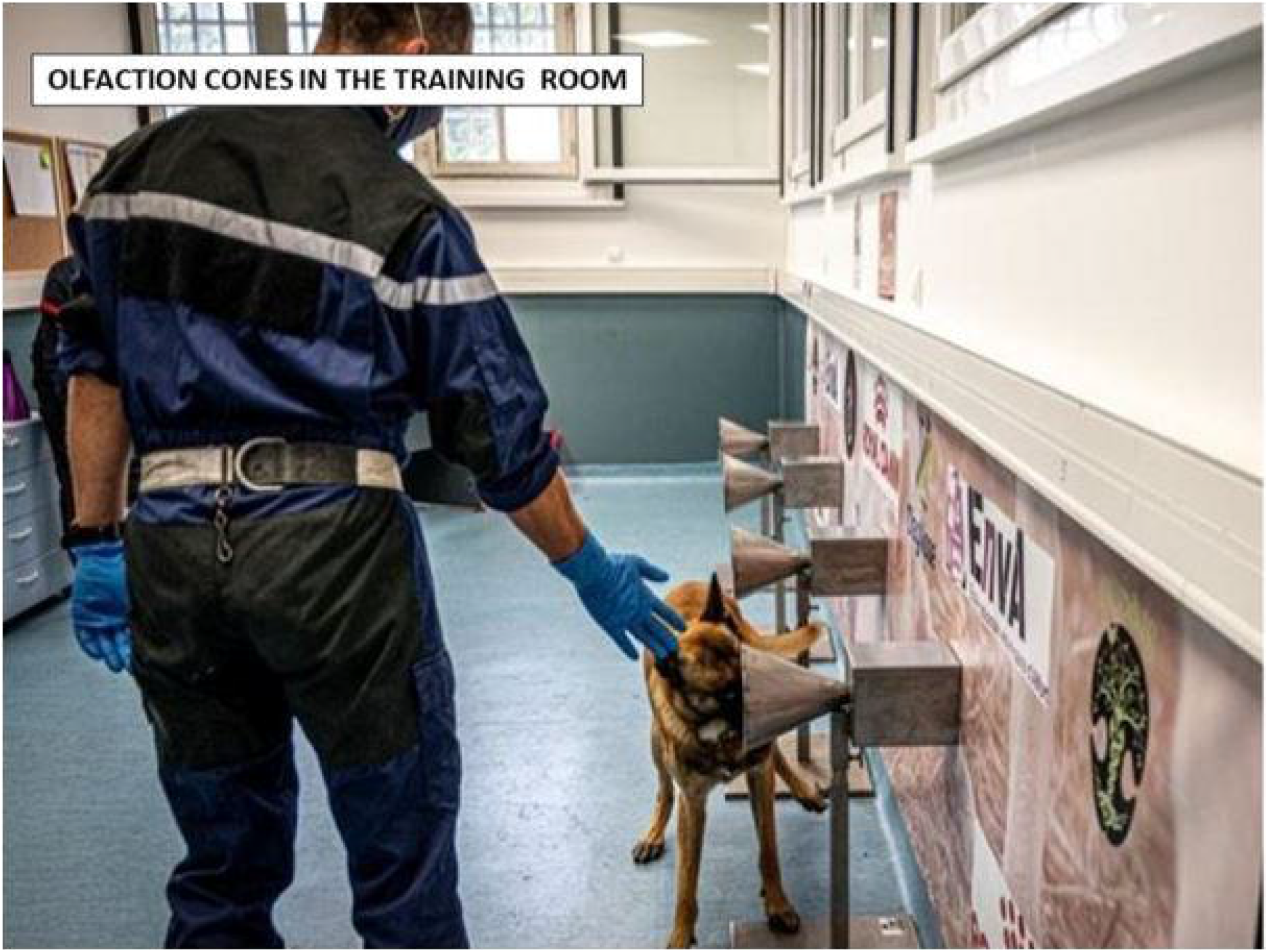
Process involving olfaction cones and dog

**Figure 1D:**
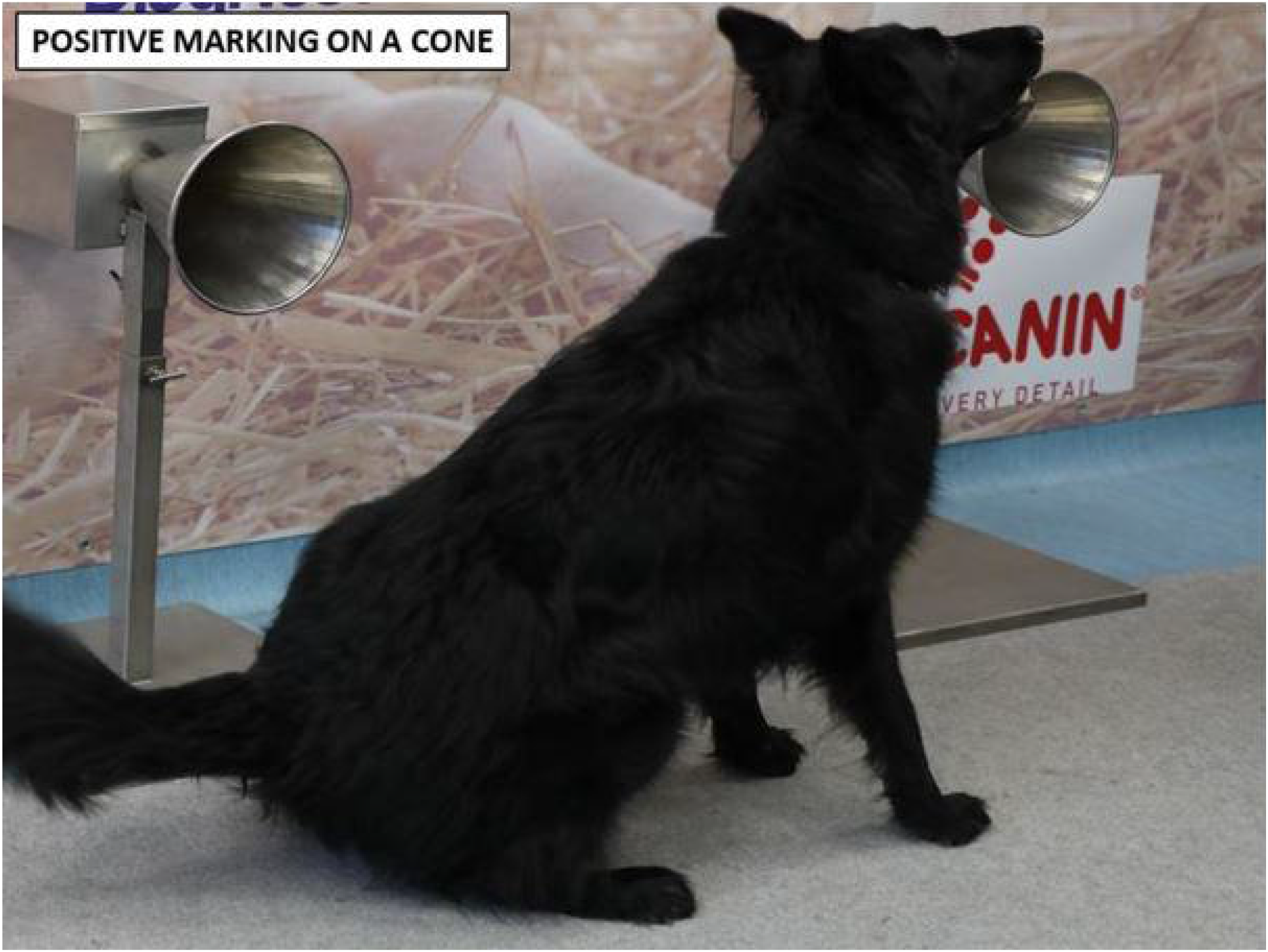
Positive marking by a dog, sitting in front of a cone containing a positive sample

### Nasopharyngeal and saliva RT-PCR

For nasopharyngeal and saliva samples, nucleic acid extraction was performed with the MGIEasy Nucleic Acid Extraction Kit (MGI Tech Co, Shenzhen, China) on an MGISP-960 instrument (MGI Tech Co). SARS CoV-2 RNA amplification was performed using the TaqPath(tm) COVID-19 CE IVD RT PCR Kit (Thermo Fisher Scientific, Coutaboeuf, France). This technique provides results expressed as a cycle threshold (Ct) for each gene target (ORF1ab, N and S genes) [6].

### Detection of other pathogens in saliva

The detection of other viruses (adenovirus, coronaviruses (229E, HKU1, NL63, and OC43), human metapneumovirus A and B, influenza, influenza A H1, influenza A H3, influenza A H1N1/2009, influenza B, parainfluenza viruses (1, 2, 3, and 4), rhinovirus/enterovirus, respiratory syncytial virus A and B) and intracellular bacteria (*Bordetella pertussis, Chlamydophila pneumoniae, and Mycoplasma pneumoniae*) was performed in saliva samples with the multiplex test QIAstat-Dx® Respiratory SARS-CoV-2 Panel QIAStat (Qiagen, Courtaboeuf, France). Briefly, 100 μL of saliva was mixed with 400 µL of NeuMoDx Viral Lysis Buffer (NeuMoDx Molecular, Inc., Ann Arbor, MI). Then 300 µl of the mix was immediately added to the SARS-CoV-2 Panel cartridge placed into the QIAstat Dx Analyzer System (Qiagen). The QIAstat-Dx Analyzer software processes controls, interprets the sample data and provides Ct values for detected targets.

### Statistical analysis

Sample size was calculated assuming that the sensitivity of the index tests was greater than or equal to 60%. To allow sufficient precision (± 10%), 93 subjects with positive nasopharyngeal RT-PCR results were needed in each of the two subgroups (symptomatic and asymptomatic participants). To account for samples excluded for technical reasons, a sample size of 110 subjects with positive nasopharyngeal RT-PCR results was needed in each of these subgroups. As preliminary results indicate that viral loads were not different between symptomatic and asymptomatic patients, the scientific committee of the study, during a planned meeting on December 16, 2020, recommended performing the analysis as soon as 93 subjects with positive nasopharyngeal RT-PCR results were included, whether symptomatic or asymptomatic.

RT-PCR results were considered positive if at least one gene was detected. For the RT-PCR technique, the Ct values reported are those for the ORF1a gene, and if not amplified, for the N gene (and for the S gene if the N gene was not amplified).

An uncertain canine detection test (the dog shows great interest in the sample, but does not immediately sit down) was considered positive. For samples that were processed by three or more dogs, only the results of two dogs were recorded at random. A sample was considered positive if both dogs marked the sample, and negative in all other cases.

Quantitative data were expressed as medians [interquartile range], and qualitative data as numbers (percentages). The diagnostic accuracy of the index tests was evaluated by calculating sensitivity and specificity. Confidence intervals were calculated by the exact binomial method. Subgroup analyses were performed according to: i) the presence of symptoms on the day of testing, ii) the Ct value of the nasopharyngeal RT-PCR, expressed as low (at least one of the 3 targets with Ct ≤ 28, i.e. high viral shedding) or high (all 3 targets with Ct > 28, i.e. low viral shedding), iii) the consumption of alcohol, coffee, food and smoking or tooth brushing before sample collection and iv) proven previous infection with SARS-CoV-2.

Sensitivity analyses were performed considering 6 alternate criteria for positivity for the reference standard: i) ≥ 2 positive targets with nasopharyngeal RT-PCR, ii) ≥ 1 positive target with nasopharyngeal RT-PCR and at least one of the 3 targets with Ct < 32, iii) ≥ 1 positive target with saliva RT-PCR, iv) ≥ 1 positive target with either nasopharyngeal or saliva RT-PCR, v) ≥ 1 positive target with either nasopharyngeal or saliva RT-PCR and at least one of the 3 targets with Ct < 32 and vi) nasopharyngeal antigen test.

Quantitative variables were compared with Wilcoxon’s test, with a significance level of 5%. The sensitivity and specificity of canine detection and nasopharyngeal antigen tests were compared with McNemar’s test. The statistical analysis was performed using R software (http://cran.r-project.org/). The reporting of results followed the Standards for Reporting Diagnostic accuracy studies (STARD 2015) guideline [7].

## RESULTS

The flow of participants is described in Figure 2. Between March 16 and April 9, 2021, 516 participants were included in SALICOV-APHP study, 403 of whom agreed to provide an axillary sample. Among them, 68 were excluded; 335 patients were thus analyzed. The proportions of females and males were similar and median age of study participants was 35 years [25-49] (Table 1). The reasons expressed by the individuals to be tested were the presence of symptoms (41.2%), being contact cases (45.7%) or others (travel, voluntary testing, etc., 13.1%). Among the participants, 43% presented with symptoms on the day of testing. The median time since the last contact and since the first clinical symptom was 5 days [0-7] and 2 days [1-3], respectively.

**Table 1:**
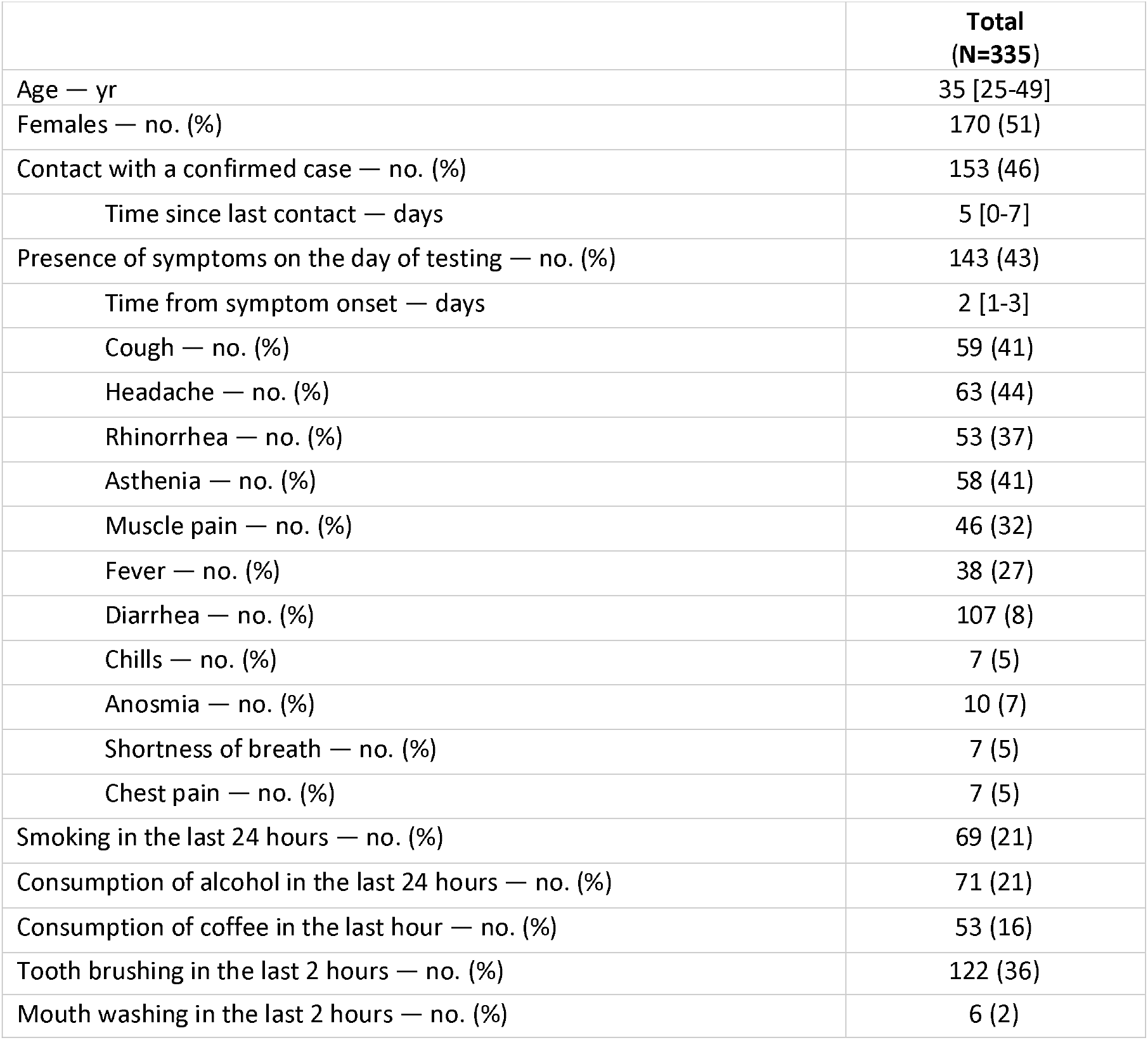
Characteristics of study participants. Results are presented as N (%) or medians [interquartile ranges].

**Figure 2:**
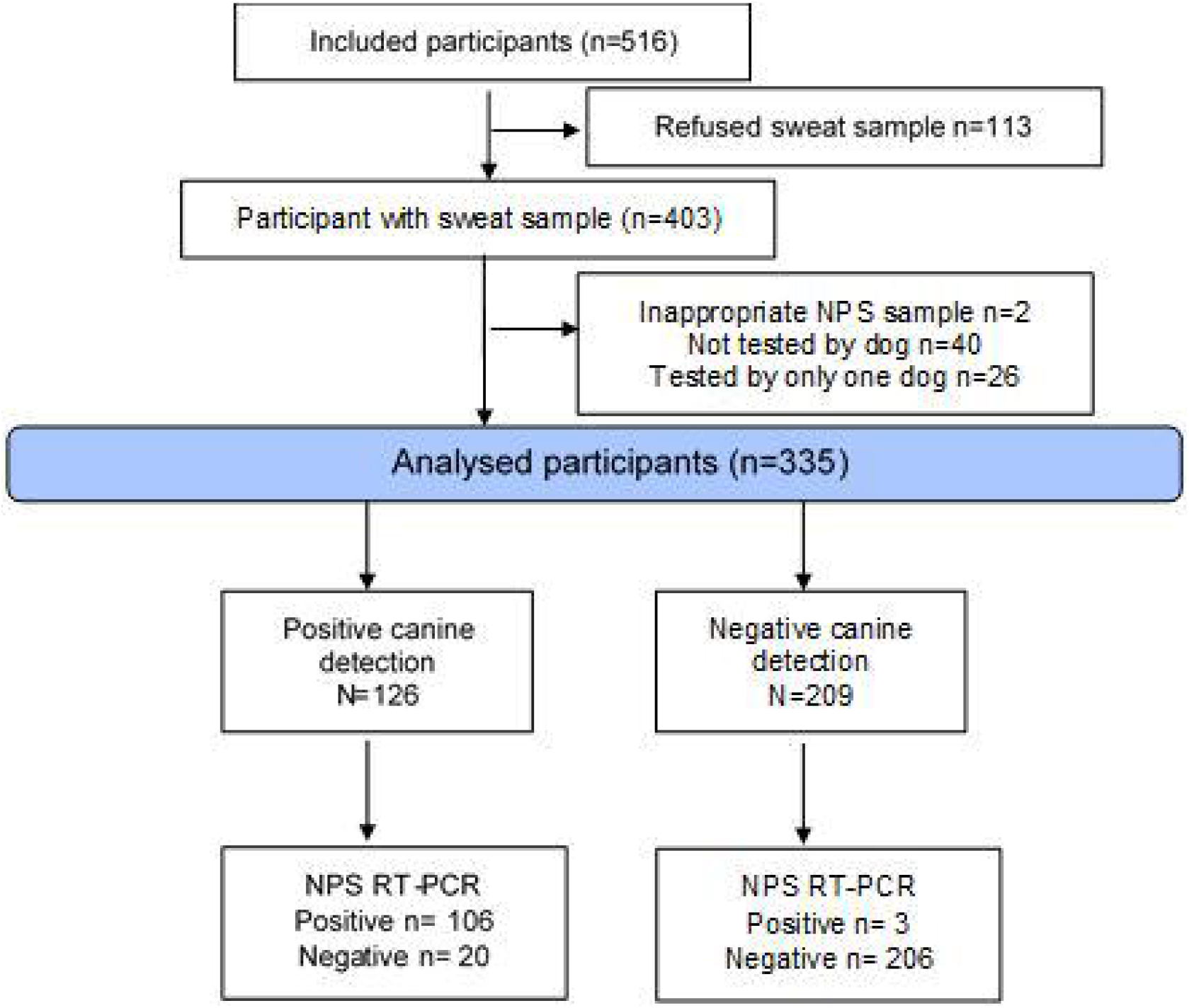
Flowchart of the study

Among the 335 participants, 109 (32.5%) tested positive on nasopharyngeal RT-PCR. The sensitivities and specificities for all the participants and according to the presence/absence of symptoms are presented in Table 2. The overall sensitivity of canine detection was 97% (95% CI, 92 to 99), reaching 100% (95% CI: 89-100) for asymptomatic individuals. The specificity was 91% (95% CI, 87 to 95), reaching 94% (95% CI, 90 to 97) for asymptomatic individuals. The PPV and NPV (Positive and Negative Predictive Values) were 84% (95% CI, 77 to 90) and 99% (95% CI, 85 to 99), respectively. The sensitivity and specificity analysis of the diagnostic accuracy of the canine detection, as compared to other reference tests, is presented in Table 3.

**Table 2:** Diagnostic accuracy of canine detection as compared to the reference standard (nasopharyngeal RT-PCR, positivity defined as at least one target gene being detected), according to the presence of symptoms in study participants.

|  | Total, n | Positive sample, n | Sensitivity<br>(95% CI*) | Specificity<br>(95% CI*) |
| --- | --- | --- | --- | --- |
| **Overall** | 335 | 109 | 97% (92 to 99) | 91% (87 to 95) |
| **Symptoms** | 143 | 78 | 96% (89 to 99) | 83% (72 to 91) |
| **No symptom** | 192 | 31 | 100% (89 to 100) | 94% (90 to 97) |

\*95% CI: 95% Confidence Interval

**Table 3:** Sensitivity analysis of the diagnostic accuracy of canine detection, as compared to several references.

| Reference standard | Total, n | Positive samples, n | Sensitivity (95% CI*) | Specificity (95% CI) |
| --- | --- | --- | --- | --- |
| **NPS RT-PCR ≥ 2 targets** | 335 | 108 | 97% (92 to 99) | 91% (86 to 94) |
| Symptoms | 143 | 77 | 96% (89 to 99) | 82% (70 to 90) |
| No symptoms | 192 | 31 | 100% (89 to 100) | 94% (90 to 97) |
| **NPS RT-PCR ≥ 1 target and Ct value < 32** | 335 | 107 | 97% (92 to 99) | 90% (86 to 94) |
| Symptoms | 143 | 76 | 96% (89 to 99) | 81% (69 to 89) |
| No symptoms | 192 | 31 | 100% (89 to 100) | 94% (90 to 97) |
| **Saliva RT-PCR ≥ 1 target** | 320 | 80 | 90% (81 to 96) | 81% (75 to 86) |
| Symptoms | 136 | 53 | 96% (87 to 100) | 65% (54 to 75) |
| No symptoms | 184 | 27 | 78% (58 to 91) | 89% (83 to 94) |
| **NPS RT-PCR ≥ 1 target or Saliva RT-PCR ≥ 1 target** | 327 | 116 | 92% (86 to 96) | 91% (87 to 95) |
| Symptoms | 142 | 79 | 96% (89 to 99) | 84% (73 to 92) |
| No symptoms | 185 | 37 | 84% (68 to 94) | 95% (90 to 98) |
| **NPS RT-PCR ≥ 1 target or Saliva RT-PCR ≥ 1 target and Ct value < 32** | 327 | 112 | 95% (89 to 98) | 91% (87 to 95) |
| Symptoms | 142 | 78 | 96% (89 to 99) | 83% (71 to 91) |
| No symptoms | 185 | 34 | 91% (76 to 98) | 95% (90 to 98) |
| **NPS antigen** | 234 | 80 | 95% (88 to 99) | 83% (76 to 89) |
| Symptoms | 123 | 60 | 97% (88 to 100) | 71% (59 to 82) |
| No symptoms | 111 | 20 | 90% (68 to 99) | 91% (83 to 96) |

\*95% CI : 95% Confidence Interval.

Measures of diagnostic accuracy did not differ according to different subgroups: with or without a medical history of COVID, and having received medical treatment or not, tobacco, alcohol or coffee, and gender (data not shown). In addition, sensitivities were similar for high (Ct ≤ 28) and low (Ct > 28) viral loads: 97% (95% CI, 92 to 99) and 100% (95% CI, 59 to 100), respectively.

Among the 234 participants who agreed to the nasopharyngeal antigenic test, the sensitivity of canine detection was greater than that of the antigenic test (97% CI: 91 to 99 versus 84% CI: 74 to 90, p=0.006), but the specificity was lower (90% CI: 84 to 95 versus 97% CI: 93 to 99, p=0.016).

To assess if the infection with other respiratory viruses could interfere with canine olfaction, saliva samples were tested with a multiplex PCR assay. Remainning saliva samples were available for 283 out of 335 participants. Out of these 283 saliva samples, 87 saliva were positive for SARS-CoV-2 and 25 samples were positive for another pathogen : rhinovirus/enterovirus (n = 11), human coronaviruses (3 OC43 and 2 NL63), parainfluenza 3 (n = 2), bocavirus (n = 1), VRS (n = 2), bocavirus / VRS (n = 1), Influenza A H1 (n = 1), human metapneumovirus (n = 1), Bordetella pertussis (n = 1). Three co-infection with SARS-CoV-2 was identified. Out of 20 individuals tested negative for SARS-CoV-2 in NPS samples but found positive with canine olfaction, 17 samples were tested for multiplex detection in saliva. Fourteen had no other pathogene, two were positive for a coronavirus (NL63 and OC43) and one was positive for SARS-CoV-2. For the diagnosis of viral respiratory infection (other than SARS-CoV-2 infection), the sensitivity of canine detection was 20% (95% CI, 7% −41%) and the specificity was 62% (95% CI, 56% −68%).

## DISCUSSION

Our results show the excellent sensitivity of SARS-CoV-2 detection by dogs using nasopharyngeal RT-PCR as the reference for comparison. These results are consistent with the results obtained previously in proof of concepts studies using sweat in hospitalized patients [5,8–11].

To our knowledge, this study is the first one carried out prospectively in the context of SARS-CoV-2 screening and the first comparing dog detection and antigenic tests.

The results detailed showed no real difference in the sensitivity observed in the different subgroups: sensitivity is always over 95% when specificity ranges from 83% to 95%.

The results obtained by canine detection using sweat samples are comparable to those of nasopharyngeal antigenic tests. However, canine detection using sweat samples is less invasive than antigenic tests on nasopharyngeal samples. Thus, the detection of SARS-CoV-2 infection by dogs could be an alternative to antigenic tests.

Dog screening for SARS-CoV-2 seems specific for COVID-19 infection and not for viral infections in general since only 2 among the 17 saliva samples in false positive patients were positive for a virus other than SARS-CoV-2. Note, however, that in both cases, it was a coronarovirus. We do not have enough patients negative for SARS-CoV-2 and positive for other coronaviruses to know whether the canine detection is specific for SARS-CoV-2 or for all coronaroviruses.

Screening people directly, without going through sweat or saliva samples, could also be considered; the advantage would certainly be increased speed. The disadvantage is the fear of dogs exhibited by a significant number of individuals and the risk of dog contamination, which is not possible with the system we have used. Direct detection by dogs should be further evaluated.

A limitation of non-invasive detection of SARS-CoV-2 infection by canine olfaction is the availability of trained dogs in this approach in light of the very significant needs if canine detection were to be considered as an alternative to antigenic tests. Another limitation is the need for certification of the dogs used for SARS-CoV-2 detection because of the risk of involving dogs whose diagnostic performance is inferior to those shown here. At the time of the study, there was no delta viriants detected, but there is no reason to believe that the results would have been different in the presence of the delta variants.

To conclude, the results obtained in our prospective study involving 335 individuals who presented voluntarily in one of the APHP testing centers in Paris support the use of canine olfaction as an alternative to antigenic tests. Canine testing is non-invasive and provides immediate and reliable results. As for antigenic tests, positive results must be confirmed by RT-PCR, especially for variant screening. Further studies will be focused on direct sniffing by dogs to evaluate sniffer dogs for mass pre-test in airports, harbors, railways stations, cultural activities or sporting events. Axillary sweat testing could remain useful for small population testing or for mobile units acting on local clusters as an alternative to antigenic tests.

## Data Availability

All data produced in the present study are available upon reasonable request to the authors

## Contributors

Study concept and design: D.G., C.D., S.K., J.L.G., J.M.T.; Patients and controls recruitment: L. C., C.E.; Data collection: C.G., C.J., V.R., L.D., G.A., S.D., A.G., M.M.; Analysis and interpretation of data: C.E, J.M.T., D.G.; Drafting the manuscript: D.G., J.M.T. All the authors read and approved the final version of the manuscript. The authors assume responsibility for the accuracy and completeness of the data and analyses, as well as the adherence of the trial, its analyses and this report to the protocol.

## Data sharing

A data sharing statement provided by the authors is available with the full text of this article.

## Acknowledgments

We thank Quentin MUZZIN, Caroline ERDEVEN, Didier ROISSE, Nicolas DIRN, Clément LEVERT, Erwan BRETON, Arnaud GALTAT, Fatma AL JASMI, Jamel ALHANAEE and Manea AL BLOOSHI for their support in conducting this study, Naim Bouazza and Frantz Foissac for proofreading the manuscript

